# Genetic determinants of severe COVID-19 in young Asian and Middle Eastern patients

**DOI:** 10.1101/2023.01.11.23284427

**Authors:** Beshr Abdulaziz Badla, Mohamed Samer Hanifa, Ruchi Jain, Maha El Naofal, Nour Halabi, Sawsan Yaslam, Sathishkumar Ramaswamy, Alan Taylor, Roudha Alfalasi, Shruti Shenbagam, Hamda Khansaheb, Hanan Al Suwaidi, Norbert Nowotny, Rizwana Popatia, Abdulla Al Khayat, Alawi Alsheikh-Ali, Tom Loney, Laila Mohamed AlDabal, Ahmad Abou Tayoun

## Abstract

Studies of genetic factors associated with severe COVID-19 in young adults have been limited in non-Caucasian populations. Here, we use whole exome sequencing to characterize the genetic landscape of severe COVID-19 in a well phenotyped cohort of otherwise healthy, young adults (N=55; mean age 34.1 ± SD 5.0 years) representing 16 countries in Asia, the Middle East, and North Africa. Our findings show enrichment of rare, likely deleterious missense and truncating variants in interferon-mediated and bacterial infection-susceptibility genes, when compared to control, mildly affected, or asymptomatic COVID-19 patients (N = 25), or to general populations representing Asia and the Middle East. Genetic variants tended to associate with mortality, intensive care admission, and ventilation support. Our findings confirm the association of interferon pathway genes with severe COVID-19 and highlight the importance of extending genetic studies to diverse populations given implications for pan-ethnic therapeutic and genetic screening options.

**Author Summary:** Based on the hypothesis that rare monogenic variants contribute to the severity of SARS-CoV-2 infection outcomes, we performed whole exome sequencing in young, previously healthy patients with severe COVID-19 of Asian or Middle Eastern origins. We found an enrichment of rare missense and truncating variants in immune-related genes, mainly associated with interferon pathways and susceptibility to bacterial infections, which can be therapeutic targets. Genetic findings tended to correlate with mortality, intensive care unit (ICU) admission, high dependency unit (HDU) admission, and invasive ventilation.

## Introduction

Since the start of the COVID-19 pandemic, as of the 2nd of January 2023, there have been over 651,918,402 laboratory-confirmed cases of COVID-19 and more than 6,656,601 deaths worldwide [1] Of these cases and deaths, 1,046,359 cases and 2,348 deaths have been reported in the UAE [2] Several risk factors have been associated with COVID-19 disease severity, including age, sex, smoking, ethnicity, and underlying health conditions like hypertension, diabetes, and cardiovascular disease [3]. Although variable and ranges from asymptomatic to severe, COVID-19 severe clinical presentation is relatively rare in young adults. Therefore, studies have focused on this age group to identify potential genetic determinants of severe COVID-19, although most such studies have been enriched for patients of Caucasian ancestry. Limited studies have focused on patients of other ancestries, such as those of Middle Eastern or Asian origins.

Zhang *et al*. reported deleterious mutations in 13 genes involved in type I interferon (IFN) pathway in COVID-19 patients with severe outcomes [4]. Other studies reported associations between *TLR7* and *IFNAR2* loss of function and severe COVID-19 [5,6,7]. Patients with severe COVID-19 outcomes were shown to have highly dysregulated expression of chemokine and interferon-related genes [8]. Evidence that SARS-CoV-2 causes Kawasaki-like, multi-system inflammatory disease in children with laboratory-confirmed COVID-19 [9,10,11] suggests that host variables controlling the immune system and host-virus interactions could contribute to COVID-19 etiology and pathogenesis.

We aimed to replicate these genetic associations with severe COVID-19 outcomes in young patients from generally understudied populations, in Asia and the Middle East. We, therefore, tested the hypothesis that inherent highly penetrant host genetic variants are associated with clinical outcomes of COVID-19 in an otherwise healthy and young patient population recruited in the United Arab Emirates (**Figure 1**).

**Figure 1:**
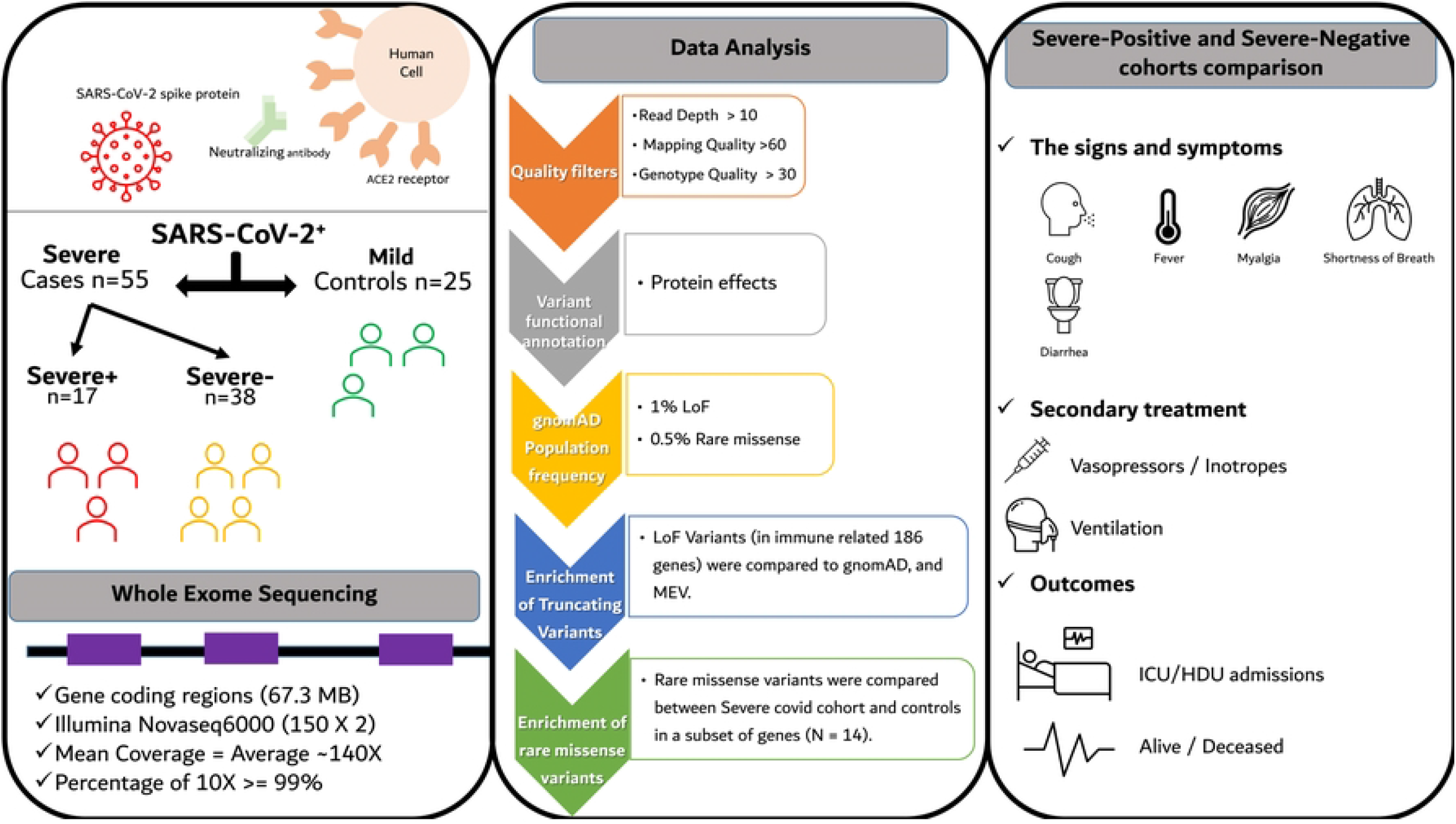
Graphical representation of study design, participants, sequencing protocol, bioinformatic analysis, and genomic and clinical characterization of patients with Severe-Covid. SARS-CoV-2; severe acute respiratory syndrome coronavirus 2; ACE2, angiotensin converting enzyme 2; LoF, Loss of function; gnomAD, Genome Aggregation Database; MEV, Middle East Variome database; ICU, Intensive Care Unit; High Dependency Unit, HDU.

## Results and Discussion

### Demographic and clinical characteristics of cohort

The cohort with severe COVID-19 consisted of 55 patients in total, of whom 83.6% were male (**Table 1**), with an average age of 34 years (range: 22 to 51 years; SD = 5.0 years) (**Table 1** and **Figure 2A**). Most patients (81.8%) were Asians while the remaining were Arabs, with an overall representation from 16 countries in Asia, the Middle East and North African region (MENA) including the UAE, Jordan, Lebanon, Syria, Palestine, Egypt, Iraq, India, Nepal, Bangladesh, Indonesia, Afghanistan, Myanmar, Pakistan, Philippines, and Ethiopia (**Figure 2B**).

**Table 1:**
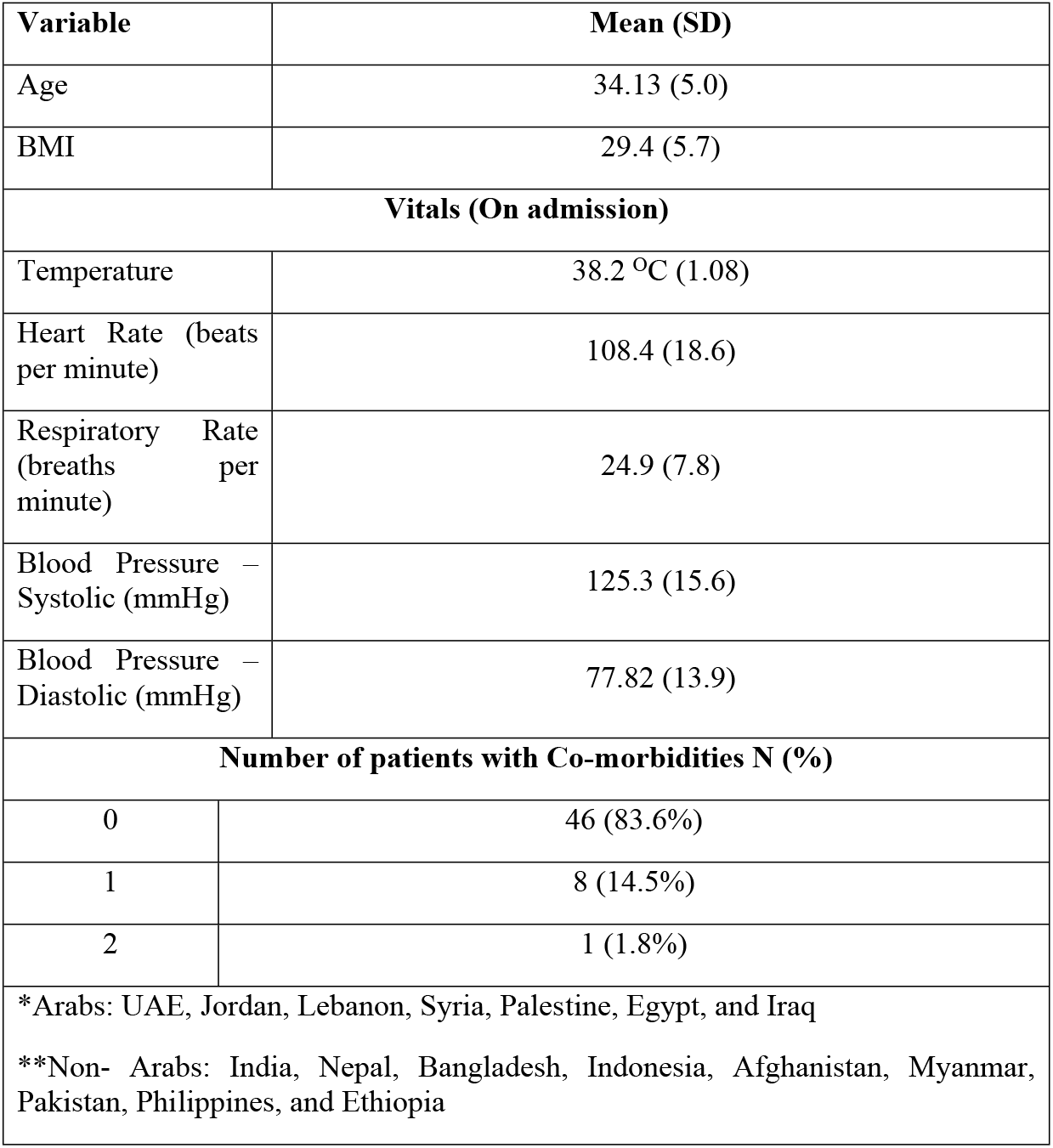
Description of Severe COVID-19 cohort (N = 55)

| **Variable** | **Mean (SD)** |
| --- | --- |
| Age | 34.13 (5.0) |
| BMI | 29.4 (5.7) |
| **Vitals (On admission)** |  |
| Temperature | 38.2 °C (1.08) |
| Heart Rate (beats per minute) | 108.4 (18.6) |
| Respiratory Rate (breaths per minute) | 24.9 (7.8) |
| Blood Pressure – Systolic (mmHg) | 125.3 (15.6) |
| Blood Pressure – Diastolic (mmHg) | 77.82 (13.9) |
| **Number of patients with Co-morbidities N (%)** |  |
| 0 | 46 (83.6%) |
| 1 | 8 (14.5%) |
| 2 | 1 (1.8%) |
| *Arabs: UAE, Jordan, Lebanon, Syria, Palestine, Egypt, and Iraq<br>**Non- Arabs: India, Nepal, Bangladesh, Indonesia, Afghanistan, Myanmar, Pakistan, Philippines, and Ethiopia |  |

**Figure 2.**
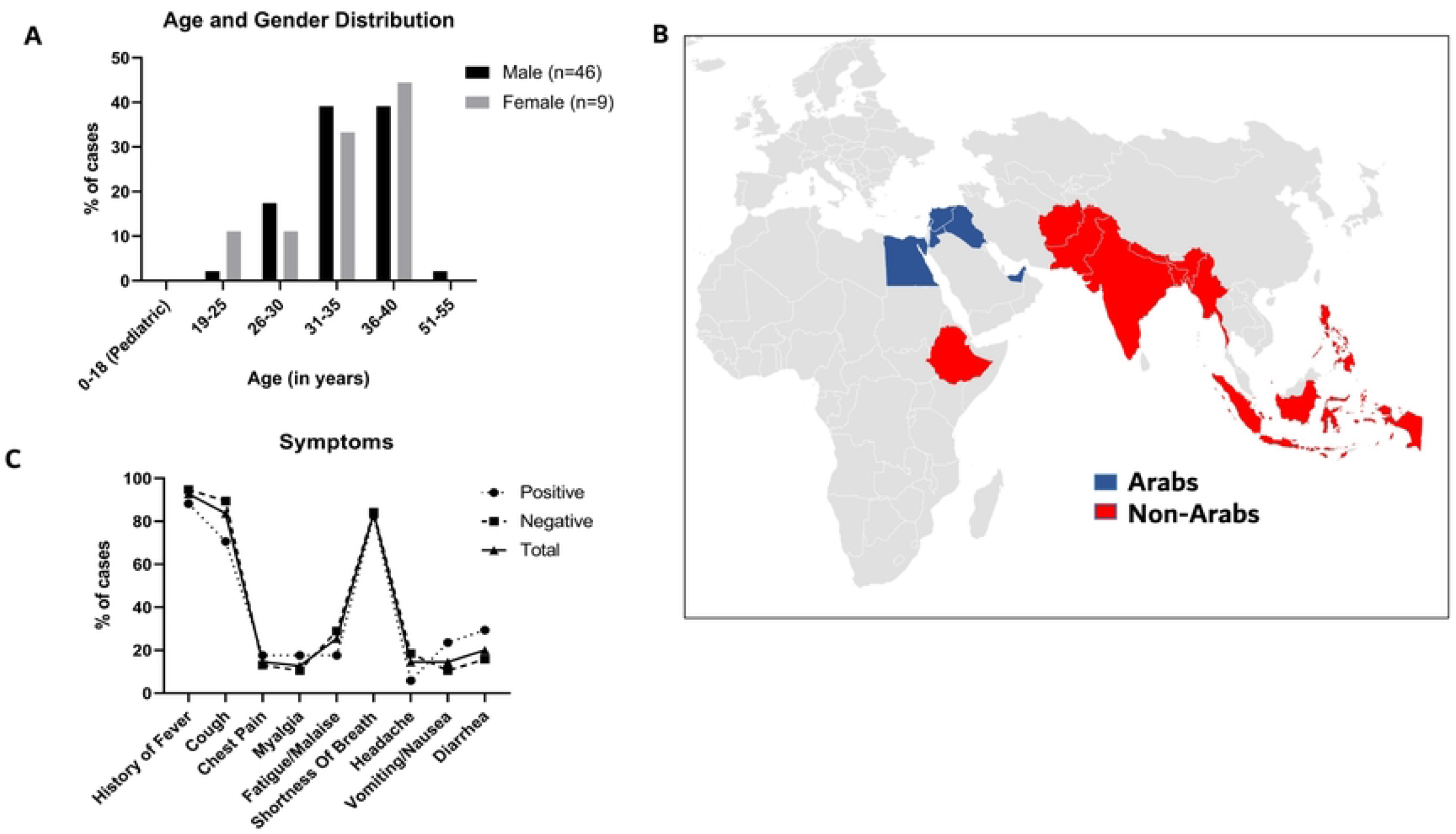
Demographics (age, gender, and country of origin) and clinical symptoms of patients with severe COVID-19. **A**. Distribution of patients age and gender, X-axis represents age (in years), and Y-axis represents percentage of cases. Grey bars represent females while black bars represent males. **B**. Geographical map representing patients’ origins or nationalities **C**. Percentage of patients with major symptoms in each grouping (Total = All patients; Positive = patients with genetic findings; Negative = patients without genetic findings). X-axis represents symptoms, and Y-axis represents percentage of cases.

Vitals were measured on admission; on average, temperature was 38.2°C (SD =1.1), heart rate was 108 beats per minute (SD = 19), respiratory rate was 25 breaths per minute (SD = 8), systolic blood pressure (BP) was 125 mmHg (SD =16), and diastolic BP was 78 mmHg (SD=14). Most patients (83.6%) had no co-morbidities (e.g., diabetes, hypertension, asthma), and 14.5% had only one co-morbidity (**Table 1**).

We recorded the patient’s symptoms on admission and throughout their stay in the hospitals (**Figure 2C**). The most prevalent symptoms within the severe cohort (N=55) were fever (92.7%), cough (83.6%), and shortness of breath (83.6%). Fatigue/malaise, diarrhea, vomiting, headache, chest pain and myalgia were observed in 25.5%, 20%, 14.5%, 14.5%, 14.5% and 12.7%, respectively. No patients had any vascular symptoms (like bleeding or lymphadenopathy), skin ulcers, conjunctivitis, or seizures.

### Genetic findings and enrichment analysis

Whole exome sequencing and targeted analysis of 186 immune-related genes (**Methods**) identified rare putative loss of function variants in 9 patients (16.4%), and rare, likely deleterious missense variants in 12 patients (21.8%). Seven patients harbored multiple rare heterozygous variants (12.7%) including one patient (COVGEN-3) with two loss of function variants in *TLR4* and *IRAK3*, and another patient (COVGEN-6) with three such truncating variants in *IFI44, TLR6 and IFNA4* (**Table 2**).

**Table 2:** Genetic Findings in young patients with severe COVID-19

| Case ID | Chromosome position | Gene(s) | Transcript | cDNA | Protein Effect | Zygosity | Effect |
| --- | --- | --- | --- | --- | --- | --- | --- |
| COVGEN-3 | chr9:120474928 | *TLR4* | NM_138554.5 | c.526_544del | p.Asn176Phefs*27 | Het | LoF |
|  | chr12:66605379 | *IRAK3* | NM_007199.3 | c.588+2T>C | p.? | Het | LoF |
| COVGEN-6 | chr1:79126263 | *IFI44* | NM_006417.5 | c.1037dupC | p.His347Serfs*4 | Het | LoF |
|  | chr4:38830867 | *TLR6* | NM_006068 | c.228_229insT | p.Glu77Ter | Het | LoF |
|  | chr9:21187471 | *IFNA4* | NM_021068 | c.60T>A | p.Cys20Ter | Het | LoF |
| COVGEN-9 | chr12:56743250 | *STAT2* | NM_005419.4 | c.1301C>G | p.Thr434Arg | Het | Rare Missense |
| COVGEN-11 | chr21:34625115 | *IFNAR2* | NM_001289125.3 | c.689T>C | p.Leu230Pro | Het | Rare Missense |
| COVGEN-14 | chr12:64890807 | *TBK1* | NM_013254.4 | c.1839G>T | p.Leu613Phe | Het | Rare Missense |
| COVGEN-18 | chr9:21206844 | *IFNA10* | NM_002171.2 | c.253C>T | p.Gln85Ter | Het | LoF |
|  | chr21:34621056 | *IFNAR2* | NM_001289125 | c.437A>G | p.Asn146Ser | Het | Rare Missense |
| COVGEN-25 | chr14:94567118 | *IFI27L1* | NM_206949.3 | c.61+1G>A | p.? | Het | LoF |
|  | chr19:4816616 | *TICAM1* | NM_182919.4 | c.1774G>A | p.Gly592Arg | Het | Rare Missense |
| COVGEN-28 | chr1:235929421 | *LYST* | NM_000081.4 | c.6079G>C | p.Val2027Leu | Het | Rare Missense |
| COVGEN-32 | chr12:56742818 | *STAT2* | NM_005419.4 | c.1466C>T | p.Pro489Leu | Het | Rare Missense |
|  | chr12:56749951 | *STAT2* | NM_005419.4 | c.250C>A | p.Gln84Lys | Het | Rare Missense |
| COVGEN-79 | chr21:34809240 | *IFNGR2* | NM_005534.4 | c.985G>T | p.Glu329Ter | Het | LoF |
| COVGEN-82 | chr1:79116171 | *IFI44* | NM_006417.5 | c.291dupA | p.Glu98Argfs*7 | Het | LoF |
|  | chr1:235929421 | *LYST* | NM_000081.4 | c.6079G>C | p.Val2027Leu | Het | Rare Missense |
| COVGEN-89 | chr19:4817579 | *TICAM1* | NM_182919.4 | c.811C>T | p.Pro271Ser | Het | Rare Missense |
| COVGEN-90 | chr1:235929421 | *LYST* | NM_000081.4 | c.6079G>C | p.Val2027Leu | Hom | Rare Missense |
| COVGEN-91 | chr2:163136505 | *IFIH1* | NM_022168.4 | c.1641+1G>C | p.? | Het | LoF |
| COVGEN-94 | chr2:163124674 | *IFIH1* | NM_022168.4 | c.2730C>A | p.Cys910Ter | Het | LoF |
|  | chr21:34713445 | *IFNAR1* | NM_000629.3 | c.341G>C | p.Trp114Ser | Het | Rare Missense |
|  | chr19:4816616 | *TICAM1* | NM_182919.4 | c.1774G>A | p.Gly592Arg | Het | Rare Missense |
| COVGEN-100 | chr9:21239446 | *IFNA14* | NM_002172.3 | c.489delC | p.Trp164Glyfs*9 | Het | LoF |
| COVGEN-127 | chr21:34707884 | *IFNAR1* | NM_000629.3 | c.131A>C | p.Asn44Thr | Het | Rare Missense |

Overall, 17 out of 55 patients with severe COVID-19 (31%; 95% CI, 20.3% - 44.0%) had at least one rare missense or truncating variant in immune-related genes and 7 (12.7%; 95% CI, 6.7% - 31.6%) had multiple such variants. On the other hand, a similar analysis on 25 patients with mild/asymptomatic COVID-19, recruited in the Middle East, identified rare variants in 3 individuals (12%) (**Supplementary Table 1**), while none of those patients had multiple rare heterozygous variants. This finding suggests that rare immunogenetic variants tend to be enriched in patients with severe COVID-19 relative to this control group (p=0.09, Two-sided Fisher’s Exact Test) (**Figures 3A** and **3B**).

**Figure 3:**
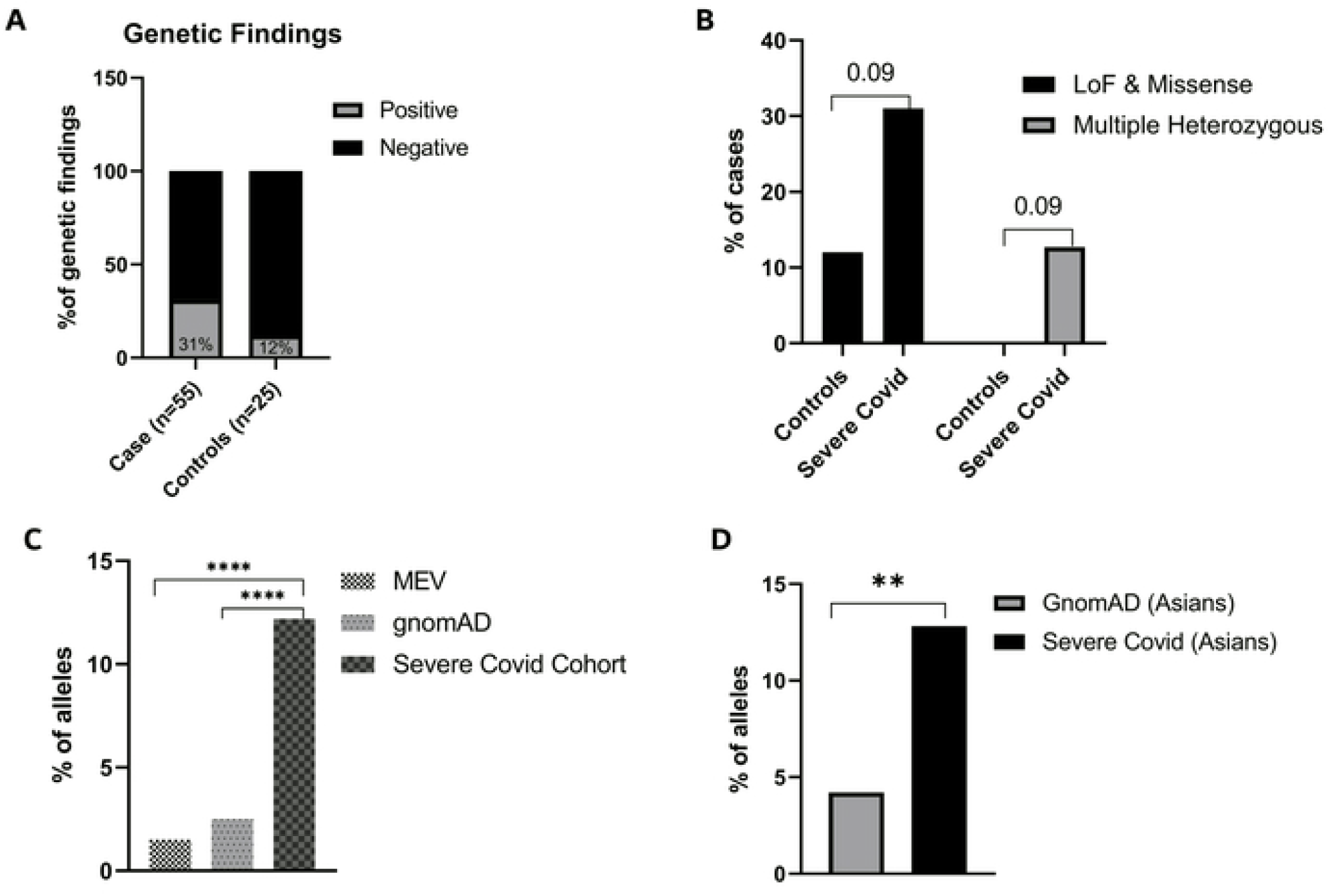
Genetic findings and burden of immune-related loss of function (LoF) and missense variants in patients with severe COVID-19. **A**. Genetic findings in cases and controls. X-axis represents cases and controls, Y-axis represents % of genetic findings. Grey filled bars represents positive genetic findings and black represents negative genetic findings. **B**. Percentage of patients or controls with at least one LoF and/or missense variants (black filled bars), or with multiple heterozygous (>1) variants (Grey filled bars). X-axis represents cases and controls; Y-axis represents percentage of each group. **C**. Frequency of truncating variants in the 10 genes identified in the severe COVID-19 cohort relative to different populations, y axis representing % of alleles. **D**. Frequency of truncating variants in Asian patients with severe COVID-19 relative to South and East Asians in gnomAD, y axis representing % of alleles. ^*^*P* < .05; ^**^*P* < .01; ^***^*P* < .001; ^****^*P* < .0001 by the Mann-Whitney test. gnomAD, the Genome Aggregation Database; MEV, Middle East Variome Database.

We then assessed whether the 10 immune-related genes (*TLR4, IRAK3, IFI44, TLR6, IFNA4, IFNA10, IFI27L1, IFNGR2, IFIH1, IFNA14*) we identified 12 truncating variants in 9 patients (**Table 2**) were similarly enriched for truncating variants in general population databases such as gnomAD and MEV database [12,13], which is a combined set of exome sequencing data from the Greater Middle East (GME) variome project (N = 1111) [14] and a Qatari cohort (N = 1005) [15]. The same immune-related genes were not significantly enriched for truncating variants in gnomAD and MEV databases (P<0.0001; Mann-Whitney test) further illustrating the enrichment of loss of function of those genes in our severe COVID-19 cohort (**Figure 3C**). Given that patients with severe COVID-19 in our study were primarily Asians (81.8%; **Figure 2A**), we performed a similar enrichment analysis focusing only on individuals of Asian origin in our severe cohort and gnomAD and found a significant enrichment of truncating variants in the Asian disease cohort (**Figure 3D**).

### Altered genes and pathways

Consistent with the critical role of IFN in viral immunity, our analyses identified several likely deleterious variants in IFN genes and related pathways (**Table 2**). There were variants either in genes coding for IFNs alpha and lambda (*IFNA4* (OMIM 147564), *IFNA10* (OMIM 147577), *IFNA14* (OMIM 147579)), IFN alpha and gamma receptors (*IFNAR1* (OMIM 107450), *IFNAR2* (OMIM 602376), *IFNGR2* (OMIM 147569), *STAT2* (OMIM 600556), *TBK1* (OMIM 604834), *TICAM1* (OMIM 607601), Toll-like receptors (*TLR4* (OMIM 603030), *TLR6* (OMIM 605403)) or IFN stimulated genes that code for IFN-induced proteins (*IFI27L1* (OMIM 611320), *IFI44* (OMIM 610468), *IFIH1* (OMIM 606951)). *IFIH1* encodes MDA5, an intracellular sensor of viral RNA that triggers the innate immune response [16], which was found to be upregulated in cell lines when infected by SARS-CoV-2 [17]. *IFI44*, was also found to be upregulated in SARS-CoV-2 infected primary normal human bronchial epithelial cells [18]. *STAT2* is critical in activating the transcription of IFN-induced genes in response to IFN stimulation. *TBK1* is a known inducer of innate antiviral type 1 IFNs [19]. *TICAM1* acts as a mediator for dsRNA-TLR3-dependent production of IFN-beta [20]. The high number of IFN pathway-related genetic findings in our severely diseased cohort highlights the potential importance of IFNs in regulating immunity and mounting an effective immune response against COVID-19.

Bacterial infection secondary to COVID-19 viral pneumonia is associated with a higher risk of death in patients with COVID-19 [21]. One of the most frequently mutated genes within our positive genetic findings group (3 patients) was the *LYST* gene (OMIM 606897) including one patient with a homozygous missense variant (**Table 2**). This gene regulates lysosomal trafficking and cytoplasmic granule synthesis, fusion, and transport [22]. In Chediak-Higashi syndrome caused by pathogenic variants in the *LYST* gene, one morbidity of this syndrome is recurrent bacterial infection(s) [22]. In addition, Toll-like receptor, *TLR4* and *TLR6*, truncating variants were found in two patients. *TLR4* is activated by lipopolysaccharides (LPS), a component of gram-negative bacteria [23], while *TLR6* is activated by diacylated lipopeptides such as lipoteichoic acid found on the cell wall of gram-positive bacteria. Alteration in genes involved in the destruction of bacteria (*LYST*) and bacterial detection by the immune system (*TLR6* and *TLR4*) could possibly increase susceptibility to bacterial infections secondary to viral pneumonia.

Among all 27 rare alleles identified in the patients with severe COVID-19, 20 alleles (74%) were in interferon-pathway-related genes, while 6 alleles (22.2%) were in bacterial infection susceptibility genes. *LYST* had the highest number of alleles (14.8%), followed by *STAT2* and *TICAM1* (11.11% each), and *IFNAR1* and *IFNAR2* (7.4% each) (**Supplementary Table 2**). The enrichment of rare variants in interferon-related genes is consistent with our previous transcriptomic analysis where we identified dysregulated interferon pathways associated with *IFIH1, IFI44, IFIT1* and *IL10* genes significantly enriched in patients with severe clinical presentation compared to mild and moderate presentations [8].

### Association of genetic variants with severe COVID-19

When combining all the positive genetic findings in the severe cohort into one group (severe-positive cohort) and comparing it to the severe group without such findings (severe-negative cohort), we found that the intensive care unit (ICU)/high dependency unit (HDU) admission rates (47.1% vs. 26.3%), mortality rate (18.8% vs. 10.5%), and the invasive ventilation support rate (23.5% vs. 10.8%) were numerically higher (almost doubled) in patients with genetic findings relative to those without (**Figure 4**). However, these three relationships did not reach statistical significance as we were most likely underpowered to find significant associations for rare genetic variants in our small cohort.

**Figure 4:**
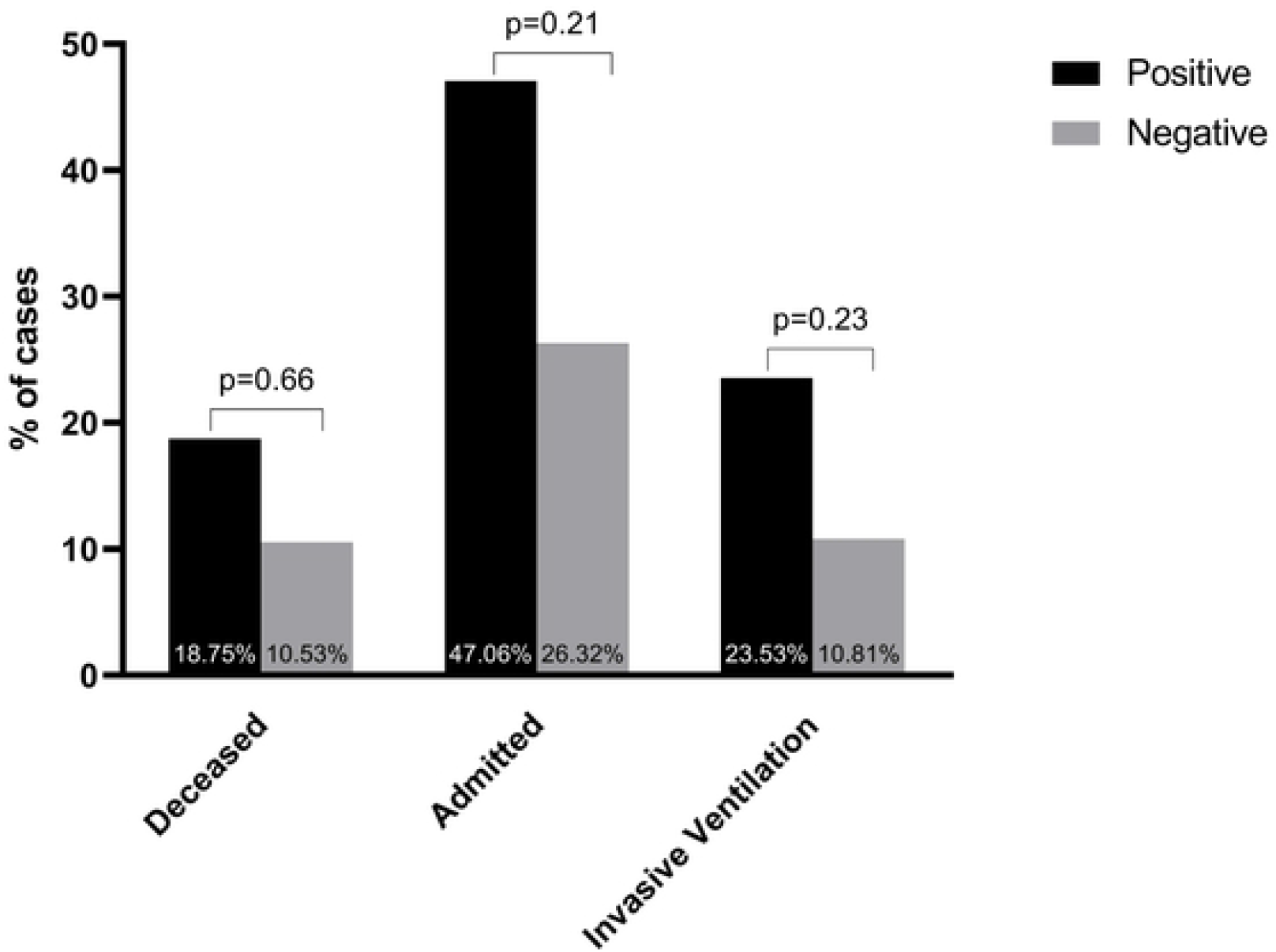
Possible association between rare genetic variants and COVID-19 severity outcomes. Mortality, intensive care unit (ICU)/ high dependency unit (HDU) admission, and ventilation support rates tended to be higher in patients with rare genetic variants (positive) relative to those without such variants (negative). However, none of those associations reached statistical significance.

### Genetic landscape of severe COVID-19 and Multisystem Inflammatory Syndrome in Children (MIS-C)

Previous work by our group found a significant enrichment of genetic variants in predominantly Middle Eastern patients with Multisystem Inflammatory Syndrome in children (MIS-C), which were associated with earlier onset of disease and resistance to treatment compared to a control group with mild SARS-CoV-2 infection [9]. The same set of immune-related genes were analyzed in the MIS-C study and this present study, identifying likely deleterious variants in 16 and 20 genes in severe COVID-19 and MIS-C, respectively. Interestingly, however, variants were overlapping in 6 genes involved in regulating Toll-like receptor signaling pathway and Interferon signaling pathways (*IRAK3, TLR6, IFNA4, IFNAR2, IFI44, IFIH1*), while majority of variants were in non-overlapping genes. This finding suggests that, while the genetic determinants of MIS-C and severe COVID-19 in young patients can be slightly different, they both share similarly altered pathways converging mainly on the interferon-signaling pathway, which has recently been shown to be disrupted in another patient population with severe COVID-19 [6].

### Strengths, limitations, and generalizability

Our study included patients from genetically underrepresented populations (N = 16 countries) in Asia and the Middle East (**Table 1**), thus complementing previous genetic studies from other commonly investigated populations. Our findings confirm the genetic determinants of severe COVID-19 in young patients from different ancestries.

While we tried to limit COVID-19 risk factors in our sample, overweight patients were not excluded from our sample. Our cohort with severe COVID-19 had a mean (± SD) BMI of 29.4 ± 5.7 kg/m^2^ which has also been associated with disease severity. The relatively small sample size limited the power of our study, which may explain the lack of statistical significance in some of the correlations between findings of rare genetic variants and disease outcomes. With a larger sample size and a stricter exclusion criterion, future studies of this nature could be more sensitized and powered to find rare genetic variants associated with disease severity.

Genetic sequencing of young, previously healthy patients with bacterial infection secondary to COVID-19 pneumonia and searching for any genetic variants associated with this outcome could unearth and determine variants leading to increased susceptibility to this complication. Efficacy testing of type 1 IFN as a therapy in patients with COVID-19 or patients with mutations in IFN pathways would determine if type 1 IFN is a viable therapeutic target.

## Methods

### 1. Study design and recruitment

Young patients with severe COVID-19 infection (N = 55) were prospectively recruited between November 2020-November 2021, mainly from Rashid Hospital, Dubai Hospital, Latifa Hospital, and Al Jalila Children’s Specialty Hospital. The control group was recruited in the UAE and Jordan, as previously described [9], in accordance with the relevant laws and regulations that govern research in both countries. This study was approved by the Dubai Scientific Research Ethics Committee—Dubai Health Authority. Patients (and their guardians) provided written informed consent for their deidentified data to be used for research.

The inclusion criteria for the severe cohort were defined as: bilateral pneumonia with more than 50% of the lungs involved, dyspnea, and SPO_2_ of less than 94% on room air. All patients had evidence of SARS-CoV-2 infection by RT-PCR. Exclusion criteria were applied to all patients with another diagnosis that can affect their illness’s course or interfere with the genetic results, such as congenital heart disease, failure to thrive, or other syndromes.

Sociodemographic information of patients such as age, gender, body mass index (BMI), and nationality were collected. Patient vitals, signs and symptoms, co-morbidities, patient status (discharged or deceased), ventilation support status or if they had been admitted to the ICU or HDU were also recorded as measures of outcomes of severe COVID-19 infection. The principal exposure studied was the presence of genetic variants.

Chi-squared Tests and two-sided Fisher’s Exact Tests were used to assess the association between nominal variables. In the 2×2 tables, the Chi-squared Test was used if all cells had a number equal to or greater than 5 and Fisher’s Exact Test for cells with small values (less than 5). SPSS software was used for all the analysis (version 25).

### 2. Whole Exome Sequencing

Whole Exome Sequencing (WES) was performed at the genomic laboratory at Al Jalila Children’s Specialty Hospital on the patients who met the inclusion criteria after written consents. DNA was extracted from peripheral blood cells using standard DNA extraction protocols (Qiagen, Germany). Following fragmentation by ultra-sonication (Covaris, USA), genomic DNA was processed to generate sequencing-ready libraries of short fragments (300-400bp) using the SureSelect^XT^ kit (Agilent, USA). RNA baits targeting all coding regions were used to enrich whole-exome regions using the SureSelect Clinical Research Exome V2 kit (Agilent, USA). The enriched libraries underwent next-generation sequencing (2 × 150bp) using the SP flow cell and the NovaSeq platform (Illumina, USA) [24,25].

### 3. Bioinformatics analysis and variant filtration

Sequencing data were processed using an in-house custom-made bioinformatics pipeline to retain high-quality sequencing reads with greater than 10X coverage across all coding regions. Variants with Read Depth > 10, Genotypic Quality > 30, Mapping Quality > 60, and allele frequency <1% in gnomAD genomes and exomes were retained.

For variant filtration and prioritization, we used signature genes (N = 186) implicated in immune responses (**Supplementary Table 3**), including cellular response to cytokine, cell mediation of immunity, immune signaling pathway, and interferon signaling pathway from reported literature [4,26-28]. We used 3 filters to retain a) LoF (Loss of Function) variants with deleterious effects on RefSeq canonical transcripts in the 186 genes, b) homozygous variants across all protein-coding effects in the same genes, and c) rare missense variants in 14 genes previously associated with severe COVID-19 [22,25], with gnomAD frequency <0.5%, for downstream analysis. We applied similar criteria for Controls (N = 25) and filtered LoF, homozygous, pathogenic, and likely pathogenic variants, DMs, and rare missense variants.

### 4. Enrichment analysis

We performed enrichment analysis for the genes with LoF or truncating variants by calculating the fractions of such variants in those genes in gnomAD [12] and in MEV [13] (Middle East Variome database, created in-house by assembling sequencing data from Qatar [15], GME (Greater Middle East)) [14] and compared those fractions with those in patients with severe COVID-19. We removed low confidence variants and applied allele frequency cutoff of 1% in gnomAD. Since majority of the cases were Asians (∼80%) we did a similar comparison using South and East Asians allele frequencies from gnomAD. The aggregate LoF allele fraction was obtained by summing the total LoF allele fraction in each of the above genes (total LoF allele counts divided by maximum allele number in the database), and Fisher’s Exact t-test was performed using Graph Pad Prism v9.2.0.

Similarly, the burden of functional variants (missense or LoF: nonsense, frameshift, canonical ±1 and ±2 at the 5’ donor and 3’ acceptor splice sites) between severe COVID-19 and controls was performed by comparing the proportion of individuals with at least 1 functional allele in each group, and *p*-values were reported.

## Data Availability

All data produced in the present work are contained in the manuscript. Any additional data will be available upon request to the authors.

